# Increased ketoacidosis from co administration of glucose-lowering drugs and Metformin: a post-marketing report study

**DOI:** 10.1101/2025.10.16.25338182

**Authors:** Ruben Abagyan, Leon Valentin Richter

## Abstract

Sodium-glucose co-transporter 2 inhibitors (SGLT2i) have become widely used not only for glycemic control but also for their protective cardiovascular effects. However, concerns about adverse events including diabetic ketoacidosis and kidney damage, have gained attention, since these agents are used both as a monotherapy and in combination with metformin. In this study, we analyzed 170,000 adverse event reports submitted to the FDA Adverse Event Reporting System for diabetic ketoacidosis (DKA), kidney injury, and lethal outcomes. Reporting odds ratios (RORs) and 95% confidence intervals (CI) revealed substantial variation in adverse event profiles across the six treatments under study. Canagliflozin shows the highest numbers on kidney damage (14.6%) of all SGLT2i monotherapies. Combination with metformin reduces it slightly (14.3%), but results in significantly elevated ketoacidosis (from 15.4% to 22.3%). Empagliflozin adverse effect profile is different, the metformin combination does not change the reported risk of kidney damage, but dramatically increases ketoacidosis from 14.7% to 27.9%, the highest number of all SGLT2i’s and their combinations. Notably, the DKA risk of each the three combinations exceeded the same parameter for a corresponding SGLT2i monotherapy of metformin alone. Metformin alone, on the other, is not associated with ketoacidosis (<1.1%), but has a high reported risk of death (12.7%) and kidney damage (18.7%), while adding any SGLT2i to the regimen reduces this kidney damage level dramatically. These observations call for individualized risk-benefit assessment of a safe and effective antidiabetic treatment regimen, particularly in patients with ketoacidosis-related comorbidities.

## INTRODUCTION

Sodium-glucose co-transporter 2 inhibitors are a relatively recent class of oral antidiabetic agents that lower blood glucose levels by blocking its reabsorption in the kidneys. Dapagliflozin, empagliflozin, and canagliflozin, collectively referred to as gliflozins, initially approved for the treatment of type 2 diabetes mellitus (T2DM), were later tested and approved as a treatment for chronic kidney disease (CKD) due to the reduction of intraglomerular pressure, and heart failure due to their effect on blood volume and blood pressure^[1;2;3]^.

Although randomized clinical trials have shown the benefits of gliflozins in reducing cardiovascular and renal risk, the larger-scale post-marketing safety data present a more complex pattern^[4;5]^. In particular, adverse events such as diabetic ketoacidosis, renal injury, and unexpected reports of increased mortality have surfaced in pharmacovigilance databases, prompting closer scrutiny^[6;7]^.

Despite the expansion of the use of gliflozins, there is growing concern that they may have different risk profiles depending on patient characteristics and combinations with other antidiabetic drugs, metformin in particular. Several case reports and observational studies highlighted the elevated risk of DKA associated with the combination of SGLT2i-metformin, particularly in patients with extracellular fluid volume depletion, prolonged fasting or reduced insulin production. Furthermore, there are statistical discrepancies between the data from the controlled clinical trial and post-marketing surveillance. In this study, we analyzed data from the FDA Adverse Event Reporting System to compare the event profiles of three SGLT2 inhibitors, as well as the combinations with metformin. Using metformin monotherapy reports as a control, we investigated the occurrence of serious adverse events, with a particular focus on lethal outcomes, diabetic ketoacidosis, and kidney damage.

Our goal was to assess whether the adverse event occurrences observed in the latest postmarketing data align with the current therapeutic recommendations and practices, and to explore whether additional considerations are warranted for specific subgroups of patients.

## MATERIALS AND METHODS

### Data source and study design

We performed a retrospective pharmacovigilance analysis using data from the FDA Adverse Event Reporting System, a publicly accessible pharmacovigilance database maintained by the U.S. Food and Drug Administration^[8]^. We extracted all reports with a single administered drug of interest listed (monotherapy), or its combination with metformin from 2004 to the first quarter of 2025. Reports submitted by healthcare professionals, patients, and manufacturers were included.

Our analysis included cohorts of patients treated with sodium-glucose cotransporter 2 inhibitors (dapagliflozin, empagliflozin, canagliflozin) and several other antidiabetic agents such as metformin, sitagliptin, liraglutide, semaglutide, and pioglitazone. In addition to those monotherapy-reports, we analyzed combinations of each of the three SGLT2 inhibitors and metformin. These fixed-dose combinations are commonly prescribed and were included in our analysis as distinct cohorts. Specifically, the combinations assessed were dapagliflozin-metformin (brand name: Xigduo XR), empagliflozin-metformin (Synjardy), and canagliflozin-metformin (Invokamet). These were identified in FAERS reports based on the co-reporting of both active substances within a single case entry, ensuring accurate attribution of adverse event signals. Only cases in which the drug was labeled as the primary suspect (PS) were considered.

### Adverse event selection and classification

We focused on adverse events (AEs) that were already known to be clinically relevant or frequently observed in our FAERS analysis. These included the following: death, diabetic ketoacidosis, kidney injury (including both renal failure and acute kidney injury), blood glucose increased, weight decreased, and fungal infections. FAERS terms for related adverse effects were aggregated where appropriate (for example, ‘diabetic ketoacidosis,’ ‘euglycemic diabetic ketoacidosis,’ and ‘ketoacidosis were considered one group).

For the kidney damage adverse event category we applied a composite definition that included multiple terms from FAERS, such as renal failure, acute kidney injury, and nephropathy. This strategy of combining related adverse effects was chosen to improve the detection of renal events, recognizing potential variability in clinical reporting terminology.

This broader inclusion was designed to capture the full scope of related adverse events and to improve the accuracy and sensitivity of the outcome detection.

### Data processing and cleaning

Raw quarterly FAERS tables were downloaded from the FDA website in structured dollar-separated text format, parsed, and joined using Unix-based tools and Python scripts. Duplicate reports were removed according to FDA recommendations, retaining only the most recent case version. The drug names were harmonized using generic names, and the combination products were categorized separately. We retained only reports with one drug (for monotherapies) or two specific drugs for a fixed combination. To ensure data quality, all entries were manually inspected for inconsistencies and errors.

### Cohort selection

This study utilized publicly available data from the U.S. Food and Drug Administration Adverse Event Reporting System, covering the period the first quarter of 2004 through the first quarter of 2025. During this period, the database included approximately 30,668,520 adverse event reports, of which 16,920,131 were classified as serious cases according to the FAERS website in July 2025.

We focused on adverse event reports that listed either dapagliflozin, empagliflozin, canagliflozin, metformin, and combinations of the gliflozins with metformin, as the primary suspect drug. Reports with a longer list of medications were excluded to avoid the ambiguity of the adverse effect assignment. Those monotherapy reports included in the analysis, as well as fixed-dose combinations with metformin, resulted in a cohort of 89,859 individual case safety reports for the specified treatment only (so called mono-therapy reports), distributed as follows: 18,369 for dapagliflozin, 20,411 for empagliflozin, 14,702 for canagliflozin, 1,863 for dapagliflozin/metformin combination, 2,657 for empagliflozin/metformin combination, 1,681 for canagliflozin/metformin combination, and 30,176 for metformin, the reference mono-therapy treatment.

All included reports specified the drug or its combination as the “primary suspect” because no other drugs were listed in selected reports.

### Statistical Analysis

The adverse event data was statistically analyzed to evaluate the relative risk profiles of selected antidiabetic drugs, with a particular focus on mortality and serious side effects such as diabetic ketoacidosis and renal injury. All analyses were conducted using simple scripting counting tools applied to selection cohorts from quarterly FAERS datasets from 2004 Q1 through 2025 Q1. For drug-event associations, we computed the reporting odds ratios with 95% confidence intervals using the formulae below. Only monotherapy reports were selected to minimize confounding by co-medications.

Metformin was used as the reference comparator in all reporting odds ratio calculations. ROR values were calculated as follows:

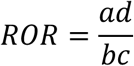

where:

a = number of reports with the drug and the event,
b = number of reports with the drug and without the event,
c = number of reports with the reference drug (Metformin) and the event,
d = number of reports with the reference drug and without the event.

Confidence intervals for the odds ratios were calculated using the Wald method based on the standard error (SE) of the log odds ratio:

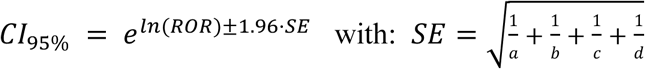

All statistical procedures, calculations, and figure generation (including forest plots) were performed using Python 3.10 (with matplotlib, pandas, and scipy libraries) and GNU Bash. Most of the RORs presented were based on a large number of reports, and the side effects with fewer reports resulted in a large CI 95%.

Significance was defined as an odds ratio whose 95% CI did not overlap with a comparator or with a ROR of 1.0 of the main comparator, metformin. All results were visualized as forest plots, with metformin value of 1 highlighted as the control.

### Ethical considerations

As the FAERS data are de-identified and publicly available, ethical review board approval was not required. The study followed FDA guidance for pharmacovigilance analyses.

## RESULTS

### Demographic analysis

Demographic characteristics of the included adverse event reports are summarized in Table 1. In six gliflozins and their combinations, males were slightly more represented in the reports, but no significant differences between the six treatments were observed. However, reports of metformin from females were more frequent. The fraction of cases with missing sex information ranged from 10.2% to 33.5%, depending on the cohort.

**Table 1.** Sex distribution of three monotherapy and three combination cases in FAERS.

| Drug name | Male, n (%) | Female, n (%) | Unknown | Total |
| --- | --- | --- | --- | --- |
| Dapagliflozin | 6,430 (35.0%) | 5,789 (31.5%) | 6,150 (33.5%) | 18,369 |
| Empagliflozin | 8,857 (43.4%) | 7,716 (37.8%) | 3,838 (18.8%) | 20,411 |
| Empagliflozin/Metformin | 1,293 (48.6%) | 1,102 (41.5%) | 262 (9.9%) | 2,657 |
| Canagliflozin | 6,257 (42.5%) | 5,480 (37.3%) | 2,965 (20.2%) | 14,702 |
| Canagliflozin/Metformin | 811 (48.2%) | 699 (41.6%) | 171 (10.2%) | 1,681 |
| Metformin | 10,290 (34.1%) | 14,343 (47.5%) | 5,543 (18.4%) | 30,176 |

### Variations of disease indications

Table 2 illustrates the distribution of the three FDA approved indications for the treatments and combinations investigated. All seven treatments were prescribed primarily for type 2 diabetes mellitus. Dapagliflozin (22.0%) and empagliflozin (15.4%) were prescribed for cardiac disorders more frequently compared to the other five treatments, while canagliflozin and metformin combinations are used more frequently in the treatment of kidney diseases. In summary, diabetes was the most frequently reported indication for all medications with available data. The differences in prescription patterns are mainly noticeable for canagliflozin, metformin, and their combination. Dapagliflozin and empagliflozin, when used as monotherapy, were often prescribed not only for diabetes but also for cardiac disorders. In contrast, combination therapies, as well as canagliflozin monotherapy, were more associated with indications related to kidney diseases than with cardiac disorders.

**Table 2.**
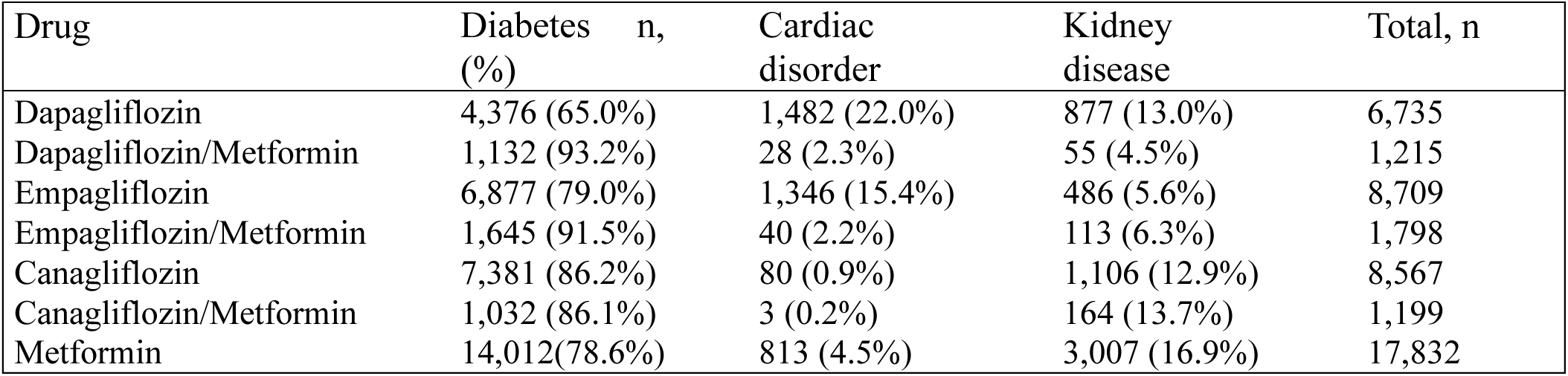
Indications of six gliflozin treatments and metformin alone in FAERS.

### Reported Occurrences and Odds Ratios for six Adverse Effect Categories

Figure 1 summarizes the frequency of six major adverse outcomes for SGLT2 inhibitor monotherapies, their metformin combinations, and metformin alone. Notably, Dapagliflozin exhibited the highest reported risk of lethal outcomes (25.71%), a high fraction of kidney injury reports (10.44%), and infections (10.49%). In contrast, the dapagliflozin/metformin combination showed a markedly lower death rate (8.32%) but a substantially higher reported frequency of diabetic ketoacidosis (20.88%) compared to the monotherapy, while being the lowest in comparison with two other combinations.

**Figure 1.**
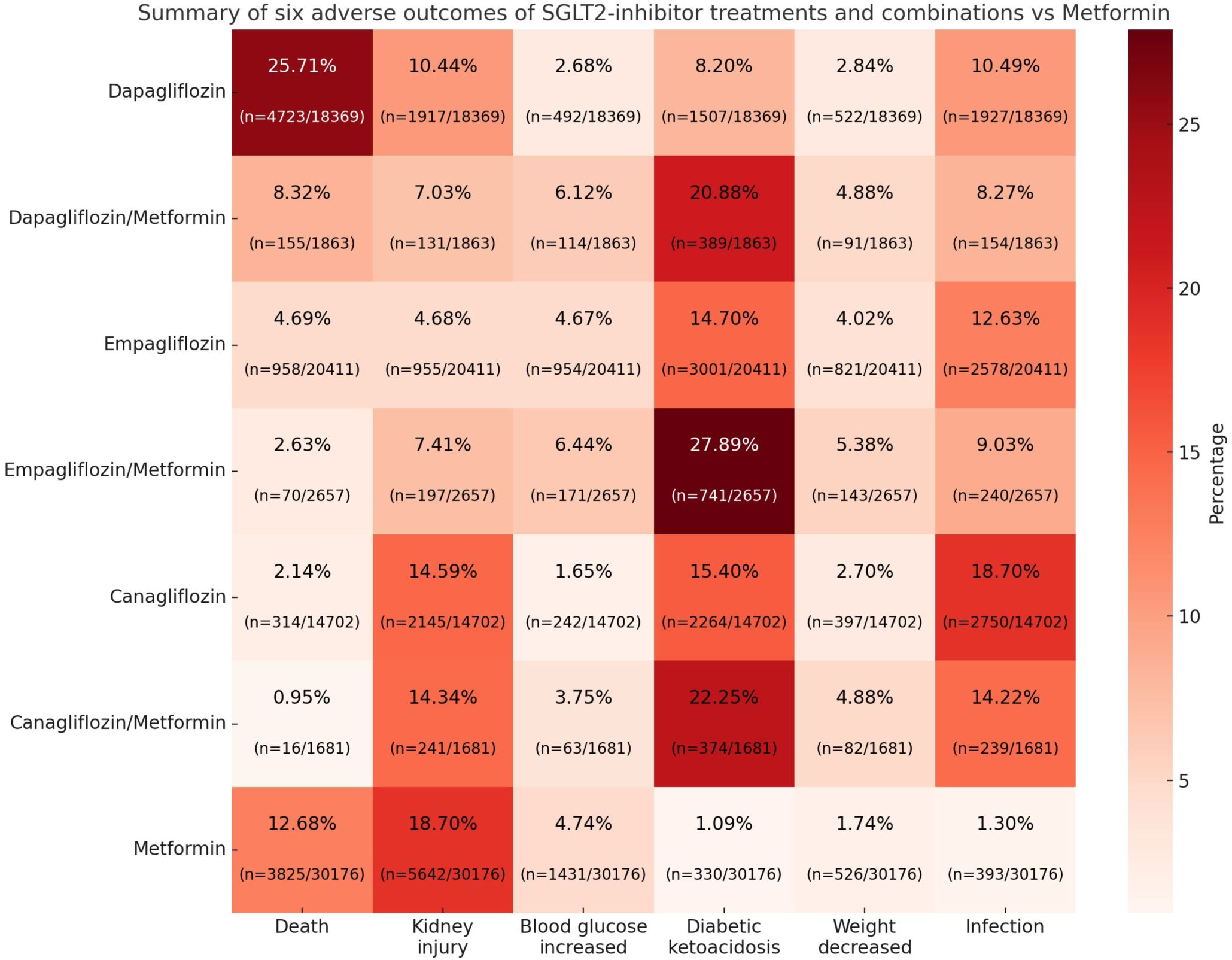
Percentage of reported adverse events for each medication.

Empagliflozin/metformin had the highest relative frequency of diabetic ketoacidosis (27.89%) among all seven treatments, while death (2.63%) and kidney injury (7.41%) remained relatively low. Canagliflozin and its combination showed the highest propensity for kidney injury (14.59% and 14.34%) among the gliflozins which was still lower than metformin alone. Canagliflozin monotherapy exhibited the highest infection rate (18.70%).

Metformin monotherapy showed the lowest frequency of diabetic ketoacidosis (1.09%) and infection (1.30%), while having the highest reported propensity for kidney injury (18.70%) and the second highest mortality rate (12.68%). The “blood glucose increased” adverse effect gave us a view into the treatment efficacy of its ability to reduce blood glucose. Although all numbers were relatively low, it should be noted that each combination with metformin increased the number of reports with this type of complaint compared to monotherapies. These observations suggest that while all three combination therapies mitigated the risk of lethal outcome, they were, at the same time, associated with an increased frequency of metabolic complications such as ketoacidosis. Surprisingly, for Dapagliflozin, the mortality reduction was counterbalanced by the increase in ketoacidosis, a great benefit of the combination treatment if the increased risk of ketoacidosis is properly managed. To evaluate statistical significance of the observed adverse effects using the reported odds ratios and their 95% confidence interface we chose metformin monotherapy as a reference, as well as performed ROR calculations for specific pairs of treatments.

### Mortality rates for SGLT2 inhibitors and their combinations with metformin

The reported odds ratios of lethal outcomes with their 95% confidence intervals attributed by the submitters to seven treatments under study, and four other widely used type 2 diabetes treatments are presented in Figure 2. Metformin monotherapy reports were used as a comparator. Dapagliflozin is associated with 2.38 times elevated reported mortality with a tight CI of [2.27,2.50] indicating its statistical significance, in comparison with metformin mono-treatment. On the other hand, canagliflozin (0.15), and empagliflozin (0.34) ROR values indicate reduced rates by factors of 6.7 and 3, respectively, with small CIs and high significance.

**Figure 2.**
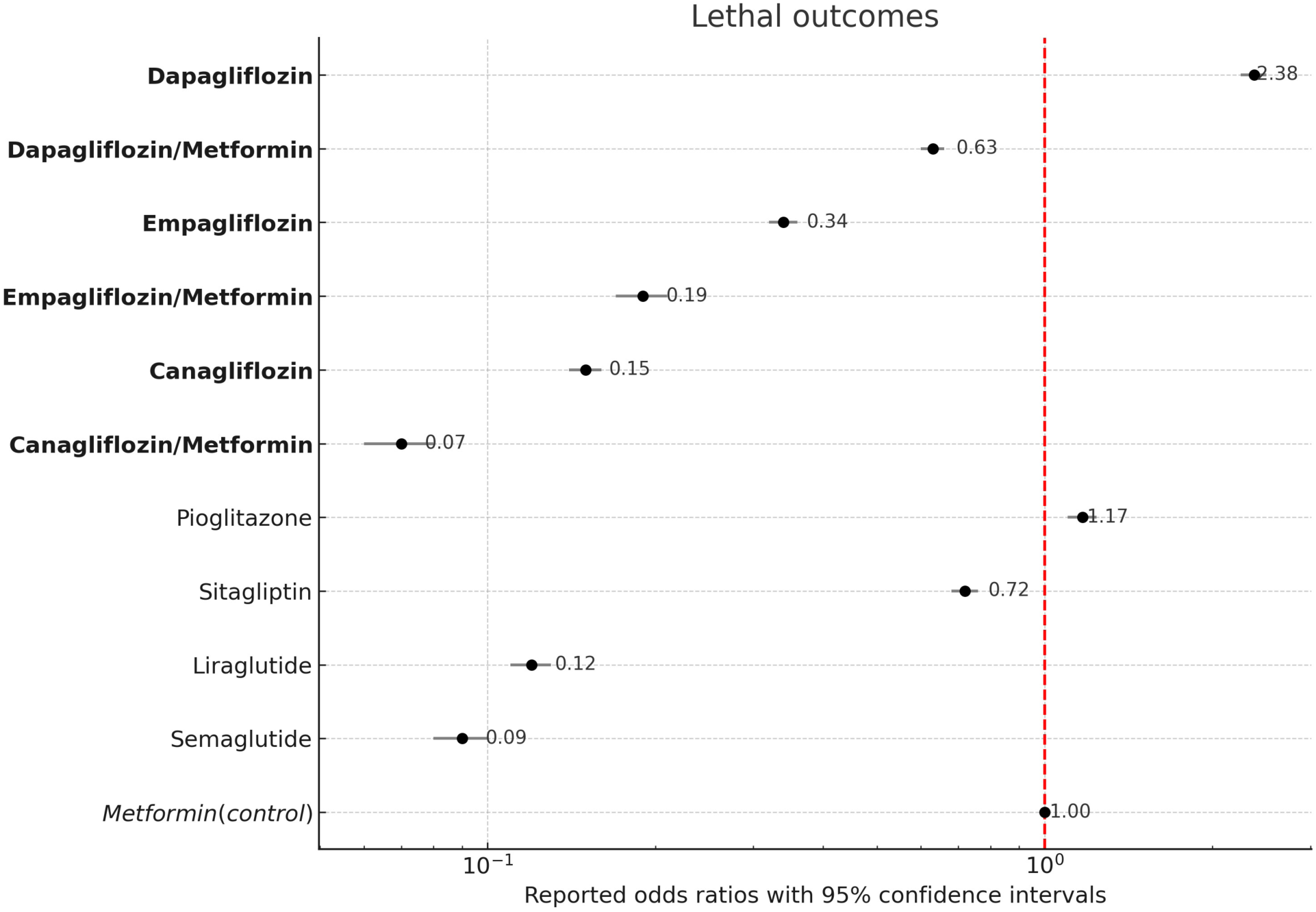
Reporting odds ratios and 95%-CIs of lethal outcomes for eleven T2DM treatments with metformin as control. The gliflozins and their combinations are ordered sequentially to highlight the synergistic reduction of mortality rates in each combination (i.e. the lethal outcome rate for each of them is lower than that of metformin alone and the corresponding gliflozin)

The most unexpected result was related to the combinations with metformin. Combining dapagliflozin with metformin reduced the mortality outcomes by an impressive factor of 3.8 (CI [3.2:4.5]) compared to dapagliflozin alone. What is even more surprising is that the ROR number for the canagliflozin-metformin combination is lower than both metformin and canagliflozin. This trend of beneficial reduced mortality effect of combinations with metformin continued for two other gliflozins but at a smaller scale. The other four T2DM treatments were comparable or lower than metformin with GLP1-agonists overlapping with metformin combinations of canagliflozin or empagliflozin. Table 3 provides a detailed overview of the odds ratios and corresponding 95% confidence intervals for reported death outcomes associated with various antidiabetic medications, using metformin as a reference. In particular, Dapagliflozin exhibited the highest mortality outcome (2.38), indicating a markedly higher risk compared to metformin alone. In contrast, agents such as canagliflozin, empagliflozin, and their metformin combinations showed substantially lower RORs, suggesting a more favorable safety profile in terms of mortality.

**Table 3.** Mortality odds ratios and 95% confidence intervals associated with antidiabetic medications compared to metformin.

| Medication | Deaths | Total reports | ROR | 95% CI (Low) | 95% CI (High) |
| --- | --- | --- | --- | --- | --- |
| Dapagliflozin | 4,723 | 18,369 | 2.38 | 2.27 | 2.50 |
| Dapagliflozin/Metformin | 155 | 1,863 | 0.63 | 0.53 | 0.74 |
| Empagliflozin | 958 | 20,411 | 0.34 | 0.32 | 0.37 |
| Empagliflozin/Metformin | 70 | 2,657 | 0.19 | 0.15 | 0.24 |
| Canagliflozin | 314 | 14,702 | 0.15 | 0.13 | 0.17 |
| Canagliflozin/Metformin | 16 | 1,681 | 0.07 | 0.04 | 0.11 |
| Pioglitazone | 1,315 | 9,048 | 1.17 | 1.09 | 1.25 |
| Sitagliptin | 1,389 | 14,750 | 0.72 | 0.67 | 0.76 |
| Liraglutide | 456 | 25,719 | 0.12 | 0.11 | 0.14 |
| Semaglutide | 390 | 31,565 | 0.09 | 0.08 | 0.10 |
| Metformin (control) | 3,825 | 30,176 | 1.00 | reference | reference |

To highlight the statistical significance of gliflozin vs gliflozin/metformin combinations, we directly evaluated ROR values and 95%-CIs for each pair, and used the combination as a control cohort. This comparison provided direct evaluation the fold reduction of mortality with both upper and lower bounds, without using metformin monotherapy as a comparator. Table 4 shows that in each pair the advantage of the combination is around sizable (two fold for empagliflozin and canagliflozin), and is statistically significant, the lowest lower bound being 1.37.

**Table 4.** Lethal outcome associated with gliflozin monotherapies vs metformin combinations.

| Medication | ROR | 95% CI |
| --- | --- | --- |
| Dapagliflozin vs. Dapagliflozin/Metformin | 3.81 | (3.23-4.51) |
| Empagliflozin vs. Empagliflozin/Metformin | 1.82 | (1.42-2.33) |
| Canagliflozin vs. Canagliflozin/Metformin | 2.27 | (1.37-3.76) |
| Dapagliflozin vs. Empagliflozin | 7.03 | (6.53-7.56) |
| Dapagliflozin vs. Canagliflozin | 15.86 | (14.11-17.82) |
| Empagliflozin vs. Canagliflozin | 2.26 | (1.98-2.57) |

\*Treatment with lower lethal outcome in blue

### Diabetic ketoacidosis reports associated with gliflozins and their combinations with metformin

Figure 3 presents the ROR values and confidence intervals for DKA across six gliflozins and their combinations and four wide-spread antidiabetic treatments in comparison with metformin monotherapy.

**Figure 3.**
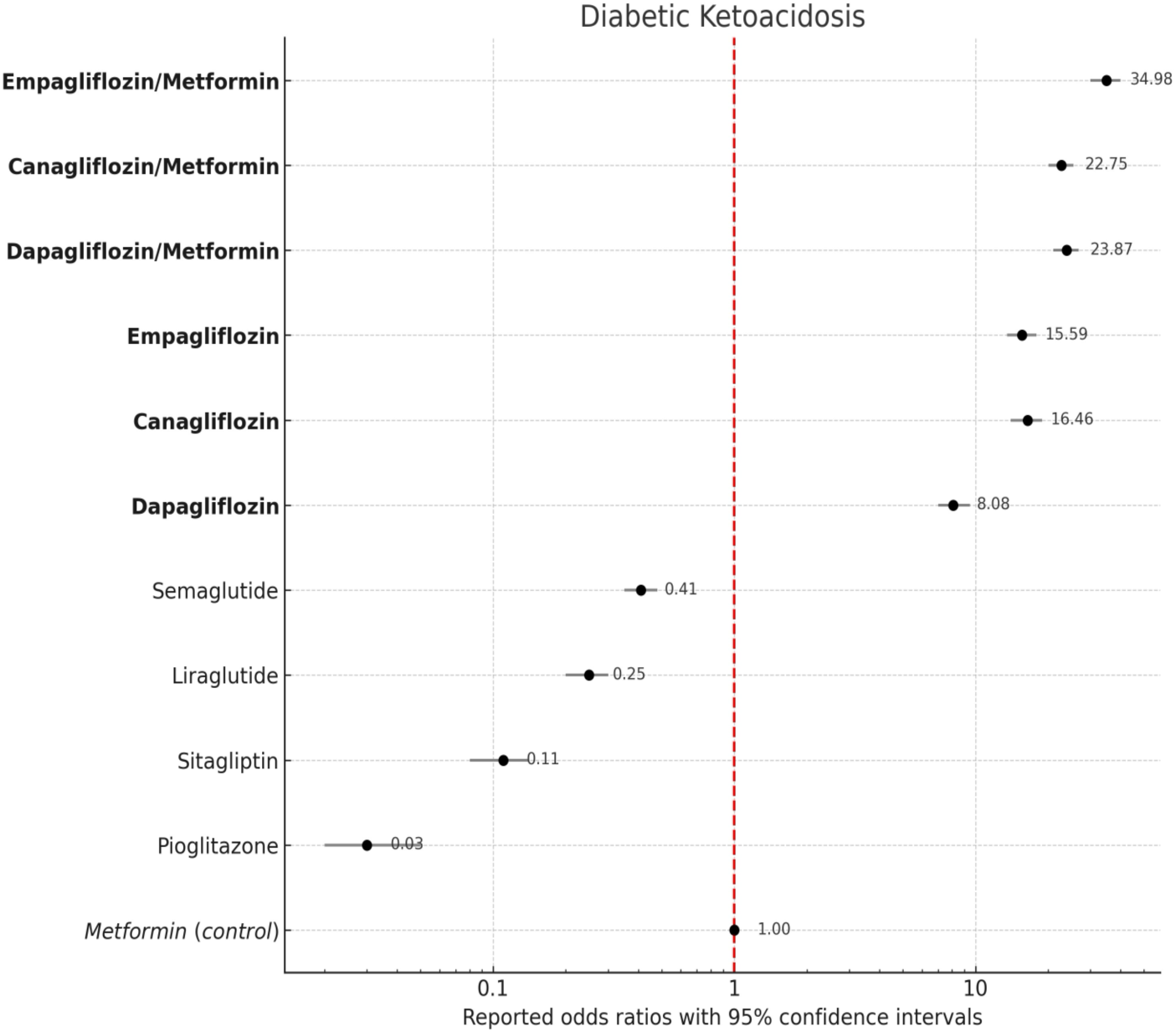
Forest plot of reported odds ratio for diabetic ketoacidosis (Metformin as control).

Notably, the highest RORs were observed for the gliflozin-metformin combination therapies, with empagliflozin/metformin (34.98), dapagliflozin/metformin (23.87), and canagliflozin/metformin (22.75) showing markedly elevated odds, with small confidence intervals, representing their statistically significant association with DKA (see the CIs in 5). The gliflozin monotherapies, on the other hand, exhibited smaller, but still alarmingly significant RORs for each of the three cases, with canagliflozin being the highest (16.46), empagliflozin (15.59) the close second, and dapagliflozin (8.08) the lowest, all in comparison with metformin alone.

In contrast, other four antidiabetic treatments under study demonstrated substantially lower risks smaller than metformin alone, namely semaglutide (0.41), liraglutide (0.25), sitagliptin (0.11), and pioglitazone (0.03). Table 5 summarizes the calculated ROR values and 95% confidence intervals for DKA associated with all ten treatments. Interestingly, while combining a gliflozin with metformin reduces the mortality, it increases ketoacidosis associated with the treatment. Further investigation is warranted to identify and confirm molecular mechanisms behind these amplified risks.

**Table 5.**
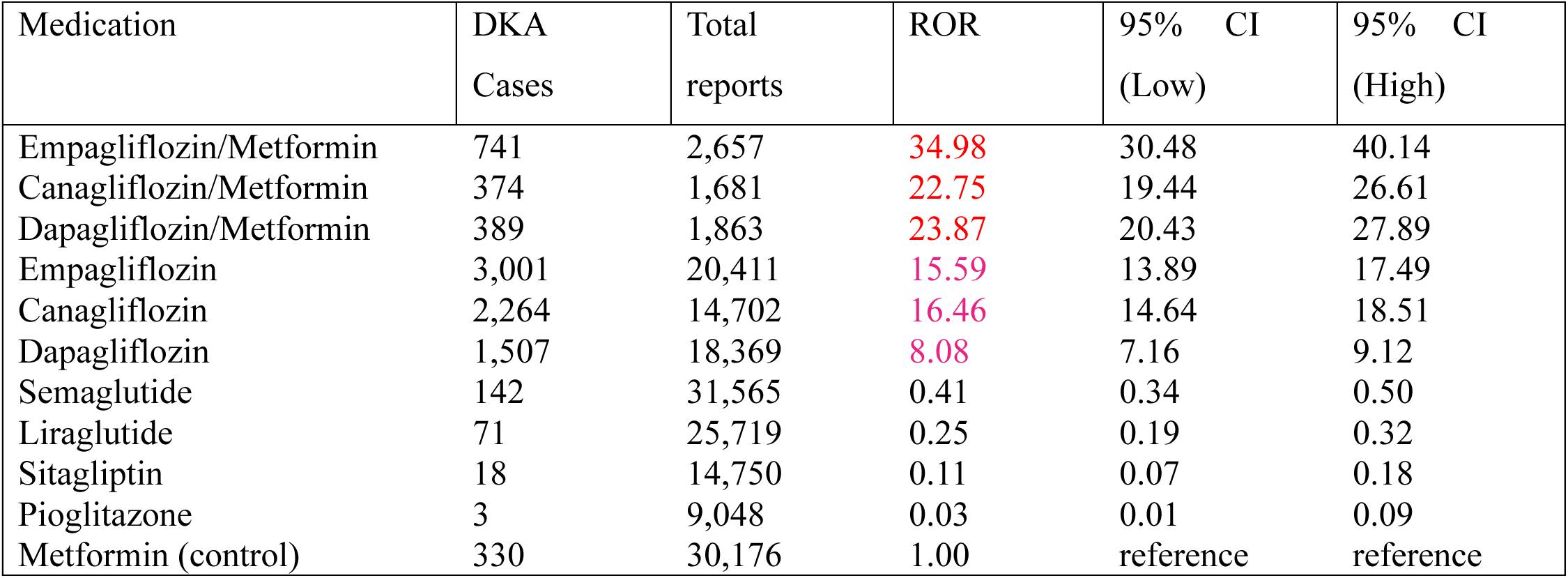
Diabetic Ketoacidosis odds ratios and 95% confidence intervals for ten treatments compared to metformin.

| Medication | DKA Cases | Total reports | ROR | 95% CI (Low) | 95% CI (High) |
| --- | --- | --- | --- | --- | --- |
| Empagliflozin/Metformin | 741 | 2,657 | 34.98 | 30.48 | 40.14 |
| Canagliflozin/Metformin | 374 | 1,681 | 22.75 | 19.44 | 26.61 |
| Dapagliflozin/Metformin | 389 | 1,863 | 23.87 | 20.43 | 27.89 |
| Empagliflozin | 3,001 | 20,411 | 15.59 | 13.89 | 17.49 |
| Canagliflozin | 2,264 | 14,702 | 16.46 | 14.64 | 18.51 |
| Dapagliflozin | 1,507 | 18,369 | 8.08 | 7.16 | 9.12 |
| Semaglutide | 142 | 31,565 | 0.41 | 0.34 | 0.50 |
| Liraglutide | 71 | 25,719 | 0.25 | 0.19 | 0.32 |
| Sitagliptin | 18 | 14,750 | 0.11 | 0.07 | 0.18 |
| Pioglitazone | 3 | 9,048 | 0.03 | 0.01 | 0.09 |
| Metformin (control) | 330 | 30,176 | 1.00 | reference | reference |

Table 6 provides a overview of the odds ratios and corresponding 95% confidence intervals for reported diabetic ketoacidosis events, associated for each pair of gliflozin and the combinations used as a control. This comparison reflect the significant increase of DKA when adding metformin to its respective gliflozin, reaching from 1.57 (canagliflozin/metformin) to 2.95 (dapagliflozin/metformin).

**Table 6.**
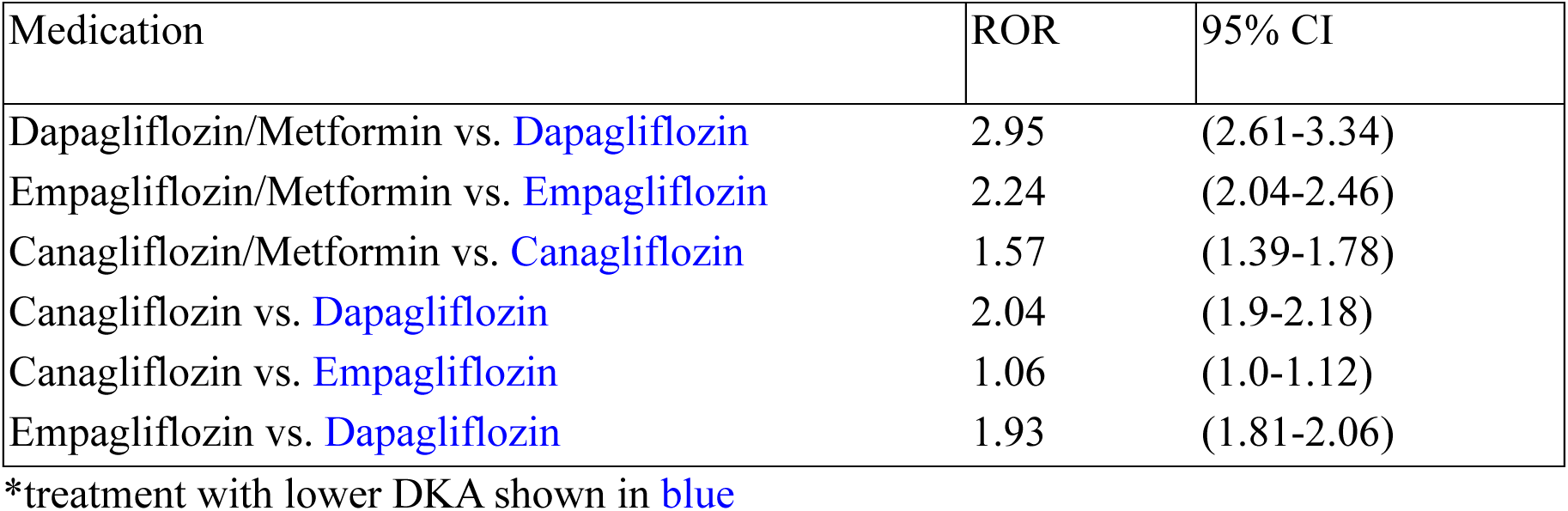
Diabetic ketoacidosis associated with gliflozin monotherapies vs metformin combinations.

| Medication | ROR | 95% CI |
| --- | --- | --- |
| Dapagliflozin/Metformin vs. Dapagliflozin | 2.95 | (2.61-3.34) |
| Empagliflozin/Metformin vs. Empagliflozin | 2.24 | (2.04-2.46) |
| Canagliflozin/Metformin vs. Canagliflozin | 1.57 | (1.39-1.78) |
| Canagliflozin vs. Dapagliflozin | 2.04 | (1.9-2.18) |
| Canagliflozin vs. Empagliflozin | 1.06 | (1.0-1.12) |
| Empagliflozin vs. Dapagliflozin | 1.93 | (1.81-2.06) |

\*treatment with lower DKA shown in blue

### Kidney damage associations with gliflozin treatments and their metformin combinations

Figure 4 displays the reporting odds ratios of kidney damage reports for ten antidiabetic therapies including gliflozins and their metformin combinations. Our control monotherapy, metformin, manifested the highest fraction of kidney damage reports of 18.4% which was somewhat unexpected given its wide use. However, the FDA revised its warnings associated with metformin use in certain patients with reduced kidney function in 2016^[9]^. Among three SGLT2 inhibitors, canagliflozin and dapagliflozin showed moderate associations with reported kidney damage (0.74 and 0.51, respectively), while empagliflozin had a significantly lower ROR of 0.21.

**Figure 4.**
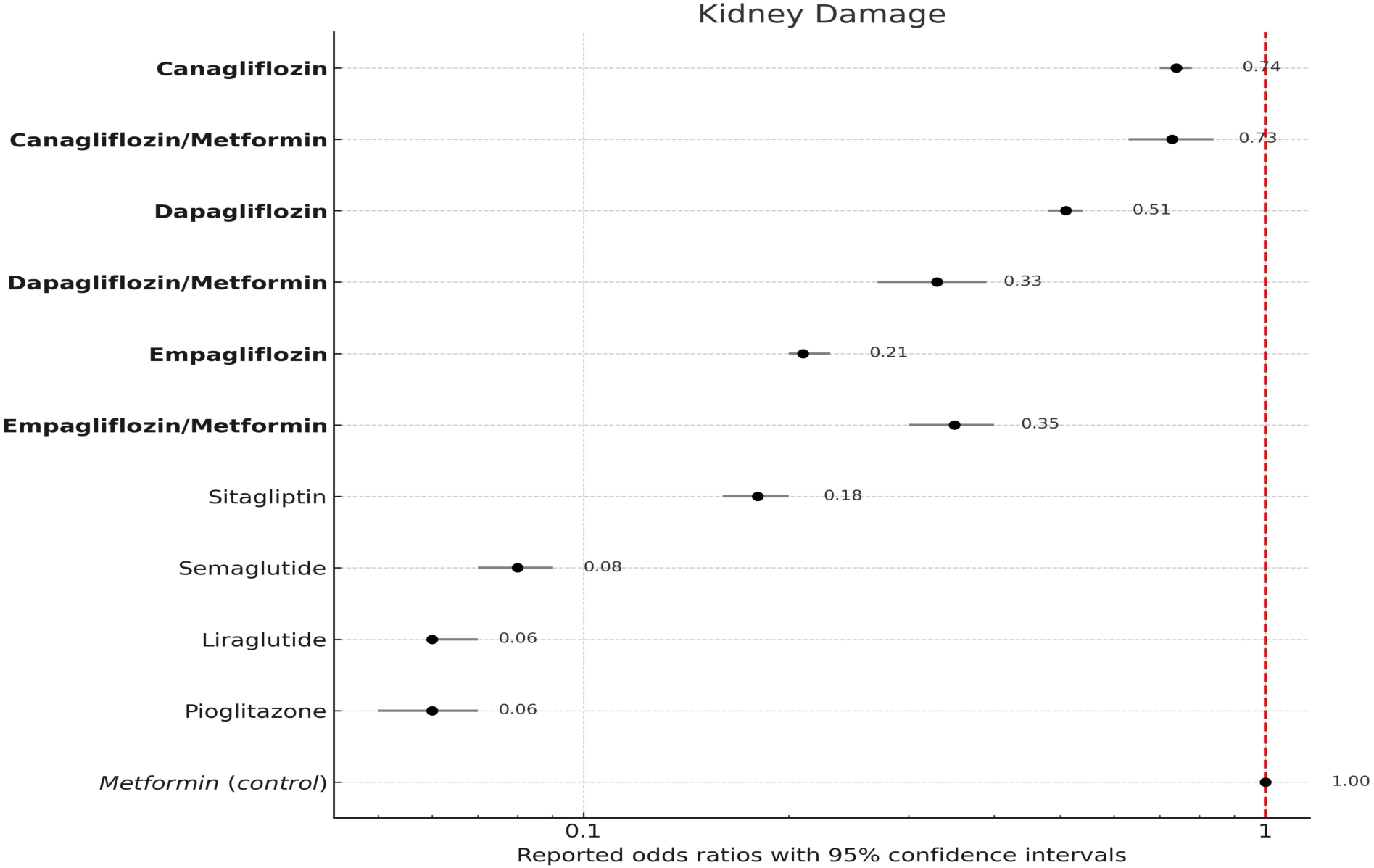
Forest plot of odds ratio for kidney damage (Metformin as control).

All evaluated SGLT2i/metformin combinations, except for empagliflozin/metformin, demonstrated lower or comparable reported odds of kidney damage compared to their respective monotherapies and the reference metformin. In particular, dapagliflozin/metformin (RORs of 0.33, vs 0.51 and 1.) and canagliflozin/metformin (RORs of 0.73 vs 0.74 and 1.) showed reductions compared to metformin. For the empagliflozin/metformin combination the ROR value increases significantly compared for empagliflozin alone (0.35 vs 0.21 and 1.) with the 95% CIs taken into considerations. This trend for empagliflozin combination is in line with the DKA side effect, and is opposite to the trend observed for the lethal outcomes.

Table 7 shows both RORs and corresponding 95% confidence intervals for all treatments under study. The lowest RORs overall were observed for pioglitazone and liraglutide, followed by semaglutide. These agents demonstrated a minimal association with reported kidney damage likely due to a different mechanism of glucose control.

**Table 7.** Kidney damage odds ratios and 95% confidence intervals associated with antidiabetic medications compared to metformin.

| Medication | KD Cases | Total reports | ROR | 95% CI (Low) | 95% CI (High) |
| --- | --- | --- | --- | --- | --- |
| Canagliflozin | 2,145 | 14,702 | 0.74 | 0.70 | 0.78 |
| Canagliflozin/Metformin | 241 | 1,681 | 0.73 | 0.63 | 0.84 |
| Dapagliflozin | 1,917 | 18,369 | 0.51 | 0.48 | 0.54 |
| Dapagliflozin/Metformin | 131 | 1,863 | 0.33 | 0.27 | 0.39 |
| Empagliflozin | 955 | 20,411 | 0.21 | 0.20 | 0.23 |
| Empagliflozin/Metformin | 197 | 2,657 | 0.35 | 0.30 | 0.40 |
| Sitagliptin | 573 | 14,750 | 0.18 | 0.16 | 0.20 |
| Semaglutide | 598 | 31,565 | 0.09 | 0.08 | 0.09 |
| Liraglutide | 356 | 25,719 | 0.06 | 0.06 | 0.07 |
| Pioglitazone | 124 | 9,048 | 0.06 | 0.05 | 0.07 |
| Metformin (control) | 5,544 | 30,176 | 1.00 | reference | reference |

Importantly, despite the comparatively higher rate of kidney damage reports of metformin monotherapy (18. 7%), most combinations of SGLT2i/metformin, except empagliflozin/metformin, particularly dapagliflozin/metformin, were associated with reduced reporting of kidney damage. These observations are consistent for dapagliflozin and, with some caveats, to canagliflozin, with a recent retrospective study of over 45,000 patients, showing that metformin co-administered with SGLT2 inhibitors significantly reduced the risk of acute kidney injury^[10]^.

For a closer focus on the kidney damage associated with dapagliflozin monotherapy, we eliminated all reports with mortality outcome from all three gliflozin cohorts and re-analyzed the kidney damage outcomes. The leading position of canagliflozin remained, with canagliflozin monotherapy showing a disproportionately higher rate of kidney-related adverse events. The order of RORs for other two SGLT2 inhibitors remained the same as well, suggesting that stronger kidney damage association observed for canagliflozin use is real and may be noticed in otherwise relatively stable patients. For a more specific evaluation of the statistical significance of the differences, we calculated the pairwise RORs and confidence intervals between individual gliflozins and their combinations with metformin (Table 8). The canagliflozin monotherapy is associated with more frequent kidney related damages compared to empagliflozin (3.48) and dapagliflozin (1.47). The empagliflozin combination with metformin does increase the kidney damage with statistical significance, but the combination outcome is more favorable compared to the metformin monotherapy (Fig. 4). These findings warrant further investigation into potential drug-specific mechanism of action leading to this adverse outcome.

**Table 8.** Kidney damage associated with gliflozin monotherapies vs metformin combinations.

| Medication | ROR | 95% CI |
| --- | --- | --- |
| Dapagliflozin vs. Dapagliflozin/Metformin | 1.54 | (1.28-1.85) |
| Empagliflozin vs. Empagliflozin/Metformin | 0.61 | (0.52-0.72) |
| Canagliflozin vs. Canagliflozin/Metformin | 1.02 | (0.88-1.18) |
| Canagliflozin vs. Dapagliflozin | 1.47 | (1.37-1.57) |
| Canagliflozin vs. Empagliflozin | 3.48 | (3.21-3.77) |
| Empagliflozin vs. Dapagliflozin | 0.42 | (0.39-0.46) |

\*An agent with improved outcome is shown in blue.

## DISCUSSION

In this retrospective study we analyzed over 170,000 monotherapy reports from the FDA Adverse Event Reporting System for three glucose lowering SGLT2 inhibitors, dapagliflozin, empagliflozin and canagliflozin, their combinations with metformin, and four other widely used antidiabetic drugs. Our primary focus was on associations between their use and the following adverse events, - diabetic ketoacidosis, kidney damage, and lethal outcomes, with a particular question about combinations of the three gliflozins with metformin, some administered in a fixed dose single pill form. We chose metformin monotherapy as a comparator for the reporting odds ratio and confidence interval calculations due its first line antidiabetic treatment status, and due its use in the gliflozin combinations. Therefore our choice of metformin as a comparator facilitated evaluation of the beneficial or harmful synergy of the two ingredients.

### Reduction of lethal outcomes in metformin combination reports

With an exception of dapagliflozin monotherapy cohort, the lethal outcome risk is not a concern for gliflozins since all numbers are lower than that of metformin, Figure 1. Dapagliflozin-related 25.71% high mortality rate can be explained, at least in part, by a particular patient demographics. In contrast to two other gliflozins and all three metformin combinations, it was prescribed not only to patients with diabetes but also ones with cardiac disorders (22.0%) and kidney diseases (13.0%) Table 2, as justified by controlled clinical trials such as DAPA-HF and DAPACKD^[1;2]^ that established cardiovascular and renal benefits. Empagliflozin, on the other hand, despite being used to treat cardiac conditions as well (15.4% Table 2) is associated with a dramatically lower mortality rate of 4.69%, Figure 1. These advantage of empagliflozin was also reported by a retrospective cohort study of patients with heart failure, in which it was associated with a lower risk of all cause mortality or hospitalization, when compared to dapagliflozin^[11]^. A surprising observation in our study was about a beneficial effect of dapagliflozin/metformin combination. Metformin, used for decades to treat diabetes type 2, exhibited the second highest mortality rate in the dataset (12.68% Figure 1). However, when dapagliflozin is combined with metformin, the mortality rate drops below mortality rates of both ingredients, even though the dapagliflozin combination-associated mortality rate is still higher than that of empagliflozin, canagliflozin, and their combinations with metformin. Overall, combining any of the three gliflozins with metformin reduced the mortality rate significantly with impressive pairwise RORs values of 3.87 (dapagliflozin) to 1.82 (empagliflozin), (Table 4, Figures 1, 2 and Table 3). This may provide a new consideration to improve safety, efficacy, and resource allocation in type 2 diabetes and heart failure management. Now let us ask ourselves if there is any downside to the metformin combinations.

### Highly elevated DKA Risk associated with SGLT2-inhibitor/metformin combinations

Diabetic ketoacidosis is a rare but potentially life-threatening complication associated with SGLT2 inhibitors according to both clinical trial data, and our analysis of the most recent postmarketing data reported here^[12]^, Figure 1.

However, the highest reporting odds ratios of DKA relative to metformin-monotherapy control were observed for the metformin combinations of the three gliflozins under study. The empagliflozin/metformin combination exhibited the highest ROR value close to 35 that corresponds to 27% vs 1% risk, followed by dapagliflozin/metformin and canagliflozin/metformin with RORs of 23.87 and 22.75 respectively. Gliflozin monotherapies are also associated with high ROR values in 8 to 17 range, but they were significantly lower than RORs for three metformin combinations, with canagliflozin vs metformin ROR of 16.46, followed by 15.59 for empagliflozin, and 8.08 for dapagliflozin, Table 5.

These DKA rates of gliflozin and their metformin combinations are substantially higher than those observed for other antidiabetic treatments, such as sitagliptin, GLP-1 receptor agonists. Our findings align with the prior pharmacovigilance studies and FDA safety alerts that reported increased risk of DKA, particularly under physiologic stress (e.g., surgery, fasting, or infection)^[3;13]^. Patients who may be particularly susceptible to the highly elevated ketoacidosis risk may fall into one of the following disease categories: type-1 diabetes with higher ketosis, poorly compliant type 2 diabetes patients, and patients with acute infections.

Notably, the metformin combination therapies are associated with greater odds of reporting diabetic ketoacidosis, and this trend was statistically significant for each of the three gliflozins as our specific pairwise comparison have shown, Table 6. The combination of dapagliflozin with metformin elevated DKA the most (ROR: 2.95 CI: 2.61-3.34).

The markedly higher risk observed with SGLT2 inhibitor combinations with Metformin may reflect cooperative non-linear effects on ketone metabolism, as metformin inhibits hepatic gluconeogenesis, potentially amplifying SGLT2-induced ketogenesis^[14]^. Clinical trials such as EMPA-REG OUTCOME and CREDENCE also reported elevated DKA risks with empagliflozin and canagliflozin used concurrently with metformin^[3;4]^.

These findings underscore the importance of patient-specific DKA risk stratification and therapy personalization for SGLT2 inhibitors combined or concurrent with metformin.

### Kidney safety benefits of gliflozins and safety variability in metformin combinations

The DAPA-CKD and CREDENCE^[3;5]^ trails established renal and cardiovascular benefits of dapagliflozin and canagliflozin in type 2 diabetes patients with chronic kidney disease. Here we used the most recent set of postmarketing reports related to these two drugs and empagliflozin, plus their metformin combinations, to investigate the kidney damage statistics. Table 7 confirms that all three gliflozins are associated with reduced kidney damage, with empagliflozin showing the lowest ROR value of 0.21 and the lowest percentage of reports of kidney damage of 4.68%

Interestingly, metformin combinations were not consistent in reported kidney damage. It stayed almost unchanged at 0.74 for canagliflozin, but showed higher kidney damage for empagliflozin combination with ROR increasing from 0.21 to 0.35. These observations reveal a more complex picture than a synergistic renoprotective effect of SGLT2 inhibitors co-administered with metformin^[10]^. For empagliflozin the combination is not beneficial with respect to kidney damage, but is beneficial for mortality, and for canagliflozin the ROR values are comparable.

This observation is in line with a comparatively higher rate of kidney damage reports for metformin monotherapy (18.7%) compared to gliflozins alone or their combinations, see Figure 4 and Table 7. The renoprotective effect of gliflozins or their combinations compared to metformin alone further illustrates kidney safety benefits of gliflozins. The other types of antidiabetic drugs we evaluated for kidney damage, including pioglitazone, liraglutide, and semaglutide (RORs 0.060.08) exhibited even lower RORs values implying their relatively safe renal profile according to the FAERS data.

In summary, these findings of variable levels of kidney damage underscore the need for individualized riskbenefit assessments in CKD patients in contrast to prescribing SGLT2 inhibitors or their combinations with metformin without distriction between gliflozins or the effect of combining them with metformin.

## CONCLUSION

In summary, the combination therapies appear to reduce the overall mortality risk, however they are associated with a significantly increased risk of diabetic ketoacidosis, which requires a careful risk-benefit assessment for individual patient profiles, before initiating such treatment combinations. Two questions remained. Is highly increased ketoacidosis the price worth paying for reduced mortality and kidney damage, and is there an additional treatment that can mitigate ketoacidosis associated with the metformin combinations, in particular? Further studies of treatment-induced changes in molecular pathways, estimated glomerular filtration rate, renal tissue inflammation and hemodynamics (see^[15]^,^[16]^) are warranted.

## DATA AVAILABILITY

The data analyzed in this study are publicly available through the U.S. Food and Drug Administration Adverse Event Reporting System (FAERS). FAERS quarterly data files for the first quarter of 2004 (2004 Q1) through the first quarter of 2025 (2025 Q1) were obtained from the official FDA repository: FAERS public dashboard. The monotherapy and combination records used in this study for statistical analyses are available from the corresponding author upon reasonable request.

## LIMITATIONS

This study based on selections from over 22 million of reports from the FDA Adverse Event Reporting System (FAERS) is subject to inherent limitations. First, the voluntary nature of submissions from healthcare professionals, consumers, and manufacturers, introduces reporting bias and can result in under-reporting or overrepresentation of certain events. Second, the quality and completeness of the reports may vary. Some characteristics, such as comorbidities, duration of therapy, and exact dosing regimens are often missing or inconsistently reported, limiting causal inference. Furthermore, FAERS lacks a defined denominator, making it difficult to calculate true incidence rates hence we rely on the reporting odds ratios with 95% confidence intervals. The disproportionality levels estimated by RORs identify statistical associations but not causality. Therefore, these findings should be interpreted as hypothesis-generating rather than confirmatory. Finally, the classification of reports as monotherapy cases is based on available entries, which may not always reflect actual clinical use due to incomplete or ambiguous drug listing. Despite these limitations, the large sample sizes and consistency of the signals observed across several types of adverse events support the robustness of our observations and underline the need for further studies to validate these findings.

## ACKNOWLEDGMENTS

We thank the Bayerische Apotheker Stiftung for funding (L.R.), Dr.Linda Awdishu, UCSD SSPPS, and Dr. Sanjay Nigam, UCSD Pediatrics for insightful discussions of molecular mechanisms and treatment practices, and Dr. Hovakim Grabski, UCSD SDSC, and Heiko Abadjian UCSD Computer Science, for computer support. We also thank Dr. Tigran Makunts for a helpful discussion of FDA pharmacovigilance practices, AE ontology, and controlled dictionaries.

## AUTHOR CONTRIBUTIONS STATEMENT

R.A. designed the computational framework and preprocessed the FAERS data. L.R. performed the data analysis and statistical calculations. Both authors contributed to rationalizing the results and writing the manuscript.

## ETHICS DECLARATION

The authors declare no competing interests.

